# Psychological aspects of caregiving after stroke: a systematic scoping review and thematic synthesis of theories

**DOI:** 10.1101/2024.10.07.24315046

**Authors:** Bethany J. Harcourt, Richard J. Brown, Audrey Bowen

## Abstract

Informal caregiving comprises a core part of stroke survivors’ rehabilitation. Providing informal care can encompass positive elements, yet adopting and sustaining this role can affect carers’ physical and mental health. Understanding carers’ experiences is important for clinical psychologists, to highlight their potential role in supporting carers’ unmet needs. Theories of stroke informal carers’ experiences exist, yet no previous attempt has been made to identify, organise and describe them, and synthesise their key themes. This PRISMA-ScR guided scoping review aimed to identify theories of stroke informal caregiving and generate new knowledge of the experience and psychological impact of caregiving, to guide stroke service improvements. Six databases were systematically searched, identifying relevant theoretical and empirical papers. Seventeen papers, presenting thirteen distinct theories, were included and the theories thematically synthesised. Two overarching themes were developed - ‘Systemic and Cultural factors’ and ‘A staged process’– containing three main themes – ‘Adjustment to aspects of post-stroke life’, ‘Emotional and psychological aspects of caregiving’, ‘Carer Needs’– and four subthemes constituting relevant stroke pathway stages. Substantial theoretical knowledge exists that was useful in generating key themes of the experiences and psychological impact of caregiving across the stroke pathway, to guide clinical psychology practice and future research directions.

## 1. Introduction

Stroke is a serious, sudden onset condition that can have unique and profound impacts on the individual. In the UK, over 100,000 strokes happen every year (SSNAP, 2023). Although strokes can happen to anyone, they do not affect everyone equally. Research has indicated that ethnic minority groups, such as people of Black or South Asian ethnicities, are disproportionately affected by stroke (Smeeton et al., 2007; Fluck et al., 2023). Furthermore, socioeconomic disparities are evident in the experience of stroke, with individuals of lower socioeconomic status experiencing stroke earlier on average than individuals from higher socioeconomic groups and experiencing a higher prevalence of post-stroke disability (Bray et al., 2018). Strokes can cause a wide range of physical, cognitive, and sensory difficulties (Törnbom, 2017; Gittins et al., 2021). People commonly need more support after experiencing a stroke (Harrison et al., 2017). A large proportion of post-stroke support is provided by informal caregivers, who may be friends, family members and loved ones (Stroke Association, 2019). Informal caregivers are a central component of the recovery and rehabilitation of the stroke survivor (Muhrodji et al., 2021; Creasy et al., 2015).

An informal carer is defined by the Stroke Association as “someone who provides unpaid help and support to family or friends” (Stroke Association, 2023, p. 1). Within the literature, these individuals are often referred to as ‘informal carers’, to enable the distinction to be made from those in paid employment as carers. The terms ‘carer’ and ‘caregiver’ are seemingly used interchangeably. ‘Carer’ and ‘caregiving’ will be used for consistency henceforth. Informal caregiving is a unique experience to each carer, potentially encompassing both challenges and positive elements (Hallam & Morris, 2014). The provision of informal care is often an integral part of the recovery and rehabilitation process for individuals who have experienced a stroke. However, research within the stroke population has mainly focussed on the experience of the stroke survivor themselves, and there is a need for more research focussed on the experience of those providing informal care or support (Harrison & Palmer, 2015).

Caregiving can have an impact on the physical health and emotional wellbeing of the individual providing care (Denham et al., 2022). Examples of this can include increased emotional and physical strain, often termed carer burden (Kerr & Smith, 2008; Achilike et al., 2020), and affected quality of life (JoCnsson et al., 2005). Furthermore, adopting the role of ‘carer’ can be a significant change to one’s identity, and for these reasons it is important that informal carers receive appropriate support to meet their individual needs (Clarke et al., 2014). Understanding the experiences of informal carers of stroke survivors is of core relevance to the profession of clinical psychology due to the role of this profession in supporting this population, as part of the wider multi-disciplinary clinical team. National Institute for Health and Care Excellence (NICE) guidelines comprise a core clinical practice guideline followed by psychological professions (British Psychological Society, BPS, 2017; Intercollegiate Stroke Working Party, 2012). Of relevance to the present study, NICE guidelines for stroke rehabilitation in adults (NICE, 2023) emphasise the importance of monitoring and supporting the needs of family members and carers, including psychological needs. This is in line with wider NICE guidance on supporting adult carers of people with health conditions (NICE, 2020).

The experience of informal caregiving from the perspective of those providing care is a valuable concept for clinical research and practice. Good Practice Guidelines encourage the involvement of carers in the development and monitoring of clinical psychology services, to ensure that interventions are high quality, accessible, inclusive, and meet their needs (Sheldon & Harding, 2010; British Psychological Society, 2017). This includes their involvement in research to develop services and interventions (British Psychological Society, 2017).

Several reviews on interventions and support needs of stroke informal carers have noted that the theoretical basis for interventions is often not reported within research (Brereton et al., 2007; Greenwood et al., 2008). Within the wider health-related literature, the enhancement of interventions through application of theory is being increasingly recognised (Campbell et al., 2000; Craig et al., 2008). The Medical Research Council (MRC) composed a framework for complex intervention development and evaluation (Campbell et al., 2000). This framework suggests research progresses through stages: relevant theory must be understood prior to developing interventions and evaluating their efficacy (Campbell et al., 2000). Complex interventions refer to those comprised of multiple, interacting components and additional dimensions of complexity (Moore et al., 2015). A wealth of empirical qualitative studies emphasise the complex nature of informal caregiving and the various interconnected elements of this (Krieger et al, 2015; Ghazzawi et al., 2016). Therefore, the MRC framework is likely of relevance to the population. Redfern et al. (2006) completed a systematic review using the MRC framework to review complex interventions in stroke care for patients, carers and family, and healthcare professionals. Findings indicated theoretical grounding to support intervention choice and development was limited, which they propose contributes to reduced intervention effectiveness (Redfern et al., 2006). In support of this, Michie and Prestwich (2010) developed and reviewed a measure to assess the extent to which behavioural interventions are theory-based. Through this work they identified numerous ways in which theory can enhance interventions, including by identifying key concepts hypothesised to be causally related to behaviour and therefore indicating appropriate intervention targets. They also identify that theory-based interventions can contribute to an understanding of the intervention’s level of effectiveness, facilitating the refinement and development of better theory (Michie & Prestwich, 2010).

In other neurological conditions such as dementia, a review of quantitative and qualitative empirical studies suggested those that were theoretically-based led to improved psychological outcomes for carers (Elvish et al., 2012). This has been reiterated in a meta-review of effective types of support for informal carers of individuals with health conditions, such as dementia, cancer and stroke (Thomas et al., 2017). Furthermore, other reviews advise that interventions should be based on theory of both the underpinning mechanism of an intervention and of the relevant context and facilitators for this to be effective, to avoid factors being overlooked that may be important in intervention implementation and effectiveness (Davies et al., 2010). Examples of factors include potential barriers to accessing support, and mediators and moderators to their effectiveness (Davies et al., 2010).

Considering these evidence and research trends in relation to clinical psychology, this profession has a core role in the support of informal carers. This can be through direct work with carers or indirectly through consultancy work to enable the wider multi-disciplinary team to provide this support (NICE, 2023; BPS, 2023). With reference to the evidenced workforce limitations within stroke services (Stroke Association, 2023), consultancy work can ensure provision of psychological support is as comprehensive and accessible as possible within services. Theories are important to clinical practice in underpinning the structure and implementation of psychological assessment, formulation and intervention (Spring, 2007; Pomerantz, 2016). This aligns with the National Stroke Service Model, which outlines both the direct and indirect role of clinical psychologists, including enabling the wider stroke team to meet the emotional and psychological needs of individuals (NHS England and NHS Improvement, 2021).

Stroke and the following experience of providing care differs from other conditions due to factors such as its sudden onset, and recovery and rehabilitation focussed trajectory. The nature of this trajectory means that some carers may provide support long-term, whereas for others, their support provision may decrease as the stroke survivor’s recovery ensues. Therefore, it is important to ascertain the theoretical understanding of the experience of providing care that is more specific to this population, as opposed to across neurological conditions more broadly. A methodological review of stroke carer research indicated that interventions would benefit from being grounded in theories that acknowledge the sudden life, role and needs changes that carers often experience (LeLaurin et al., 2019).

Where theories and conceptual frameworks have been utilised in previous studies of informal caregiving in stroke, these often pertain to stress, coping and adjustment that have been developed in other clinical populations and then applied to stroke informal carers (Lee & Song, 2022; Hussain et al., 2014; Eldred & Sykes, 2008). Examples include the Transactional model of Stress and Coping (Lazarus & Folkman, 1984), Sociocultural Stress and Coping Model for Caregivers (Aranda & Knight, 1997) and cognitive adjustment theories (Thompson, 1991). Other informal caregiving theories exist that have been developed through systematic reviews and empirical work with informal carers of stroke survivors. Some of these focus on the preparation of the individual for undertaking the care role (Lutz et al., 2017), and others focus on stages of the caregiving experience an informal carer may transition through along the stroke trajectory (Cameron & Gignac, 2008).

Previous scoping reviews have mapped the evidence regarding specific elements of the experience of informal caregiving after stroke. These include the quality of life (Moura et al., 2022; Tsiakiri et al., 2023) and the health needs (Felix et al., 2021) of carers. However, no previous attempt has been made to organise and synthesise the theoretical literature in this area. Scoping reviews help to identify, describe, and codify theoretical concepts in an area, map the theoretical underpinnings of the current understanding of a topic and link to implications for clinical practice (Peters et al., 2021; Levac et al., 2010).

This scoping review therefore aims to identify and synthesise theories of informal caregiving following stroke that pertain to the psychological impact of caregiving (e.g., the mental health and wellbeing of the individual providing care), to guide the improvement of stroke services. ‘Theories’ will be used throughout this paper to promote consistency and will refer to theoretical conceptualisations that predict or explain the experience of providing informal care to a stroke survivor and the psychological impact of this.

## 2. Method

### 2.1 Protocol pre-registration

The systematic scoping review protocol was pre-registered on the Open Science Framework Registries database (https://doi.org/10.17605/OSF.IO/9HP6M). Although our initial intention was to conduct the review across multiple neurological conditions, the extent of the theoretical literature on caregiving in stroke indicated that a more focused review was possible and appropriate. An update to the initial protocol was therefore made. The review adhered to the Joanna Briggs Institute methodology and guidance for scoping reviews (Peters et al., 2020).

### 2.2 Study eligibility

The review’s inclusion criteria were peer-reviewed publications that: (1) either proposed, described, evaluated or reviewed a singular or multiple theories around informal caregiving in stroke; (2) focused on the experience of, and processes involved in, the psychological impact of caregiving (3) were theoretical or empirical, using a qualitative, quantitative, or mixed-methods research design; (4) were available as full articles (i.e. not only as abstracts); (5) were written in the English language; (6) related to adult carers, aged 18 years and above, of people with stroke.

In these inclusion criteria, empirical papers refer to those that generated novel data through their study and conceptualised this into a theory. Theoretical papers refer to those which did not involve generation of empirical data, yet were based on reviews of existing literature, to conceptualise a theory. Studies and literature conducted in any stroke healthcare or academic setting were included.

### 2.3 Search strategy

For review comprehensiveness, a range of electronic databases were searched from the earliest date available to December 2023: MEDLINE, PsycINFO, PubMed, Embase, CINAHL Plus and Web of Science. Databases were selected based on their usage in previous scoping and systematic review studies completed around the topic of informal caregiving in stroke (Krishnan et al., 2017).

The following search terms were required to be present in the title or abstract of papers and were combined with Boolean operators: (stroke* OR cerebrovascular accident*) AND (theor* OR framework* OR conceptual*) AND (caregiv* OR care OR caring OR support* OR informal car*). Slight modifications to the search strategy were employed for the PubMed and Web of Science databases due to their requirements for lengths of search terms: the “informal car*” term was modified to “informal care*”.

Search results were imported into Rayyan and duplicates were detected and removed. The lead reviewer (BH) completed an initial screening of the titles and abstracts of papers to remove results not meeting the inclusion criteria. A second reviewer (JW) independently screened 20% of these papers at the title and abstract stage. Inter-rater reliability results were calculated, with a Cohen’s kappa value of 0.99 indicating almost perfect agreement. Full text screening was subsequently completed by BH and the second independent reviewer (JW). Inter-rater reliability calculations for the full text screening indicated a Cohen’s kappa value of 0.90, almost perfect agreement. Any disagreements of study eligibility and inclusion were resolved via discussion between reviewers, and with the other authors.

Following completion of database searches, forwards and backwards citation searches of the papers meeting the inclusion criteria were completed for validity and completeness of literature searching, and to ensure all relevant papers were identified.

### 2.3 Data extraction

Data extraction was completed by the lead reviewer, BH, for all included studies. Study characteristics were extracted including name of theory; topic or focus of the theory; author name(s); year of publication; country of publication; the type of study (i.e. empirical or theoretical) that had generated the theory; the population and population demographics in which the theory was developed or applied. Qualitative data fragments were extracted from the studies, comprising sections of the text that present relevant theoretical concepts. All data were transferred to a data extraction spreadsheet.

### 2.4 Thematic Synthesis

Following scoping review guidance (Thomas & Harden, 2008; Peters et al., 2020), thematic synthesis of the qualitative data was completed, with a view to identifying themes and concepts relating to the experience and psychological impact of caregiving in stroke. The three stages of thematic synthesis were followed (Harden et al., 2008). Firstly, the extracted qualitative data fragments describing the theory were uploaded to NVivo12 (QSR International Pty Ltd. Version 12, 2018) software for qualitative data, and inductively read to allow line-by-line free coding. In the second stage, the inductive coding of the data, codes were organised into descriptive themes. Following this, analytical themes were generated by moving beyond the descriptive theme level to create new interpretative constructs (Thomas & Harden, 2008). Throughout the thematic synthesis process, discussions within the research team aided the refinement and modification of codes and themes. An example of code and theme development can be seen in Figure 1. The resulting themes were presented both textually and visually.

**Figure 1:**
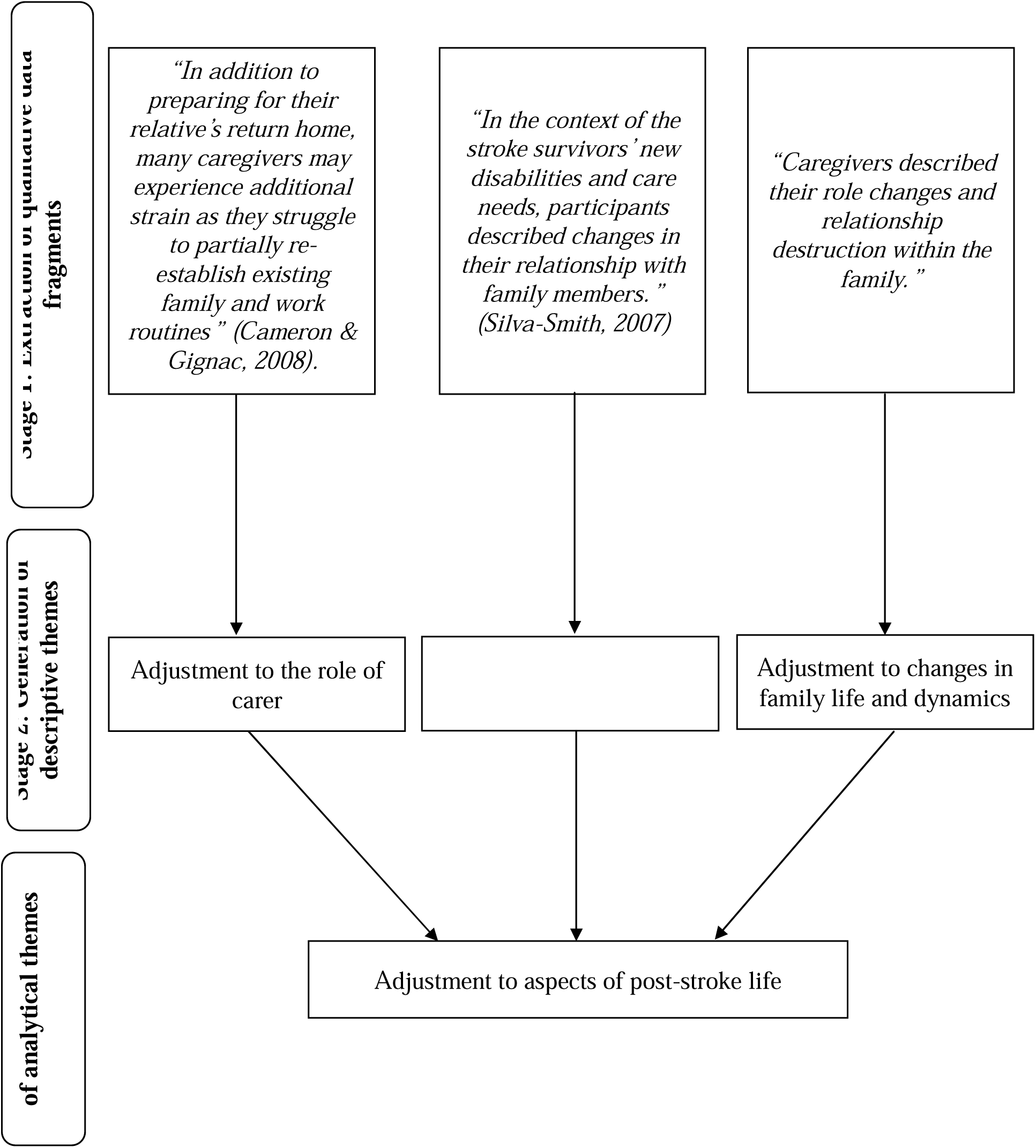
Example of thematic synthesis theme development process.

## 3. Results

### 3.1 Characteristics of included theories

Figure 2 presents an outline of the search process, based on the Preferred Reporting for Systematic Reviews and Meta-Analyses extension for Scoping Review (PRISMA-ScR) extension (Tricco et al., 2018). A total of seventeen papers were identified for inclusion in the current review, as summarised in Table 1. Studies were conducted in a variety of countries, mainly Canada and the United States. Thirteen theories were proposed in the seventeen studies, with one theory ‘Timing it Right’ reported in five papers. Most of the papers reported empirical studies. A minority of papers included stroke survivors as well as carers. There is very little information on ethnicity of participants, with papers mainly reporting nationality.

**Figure 2:**
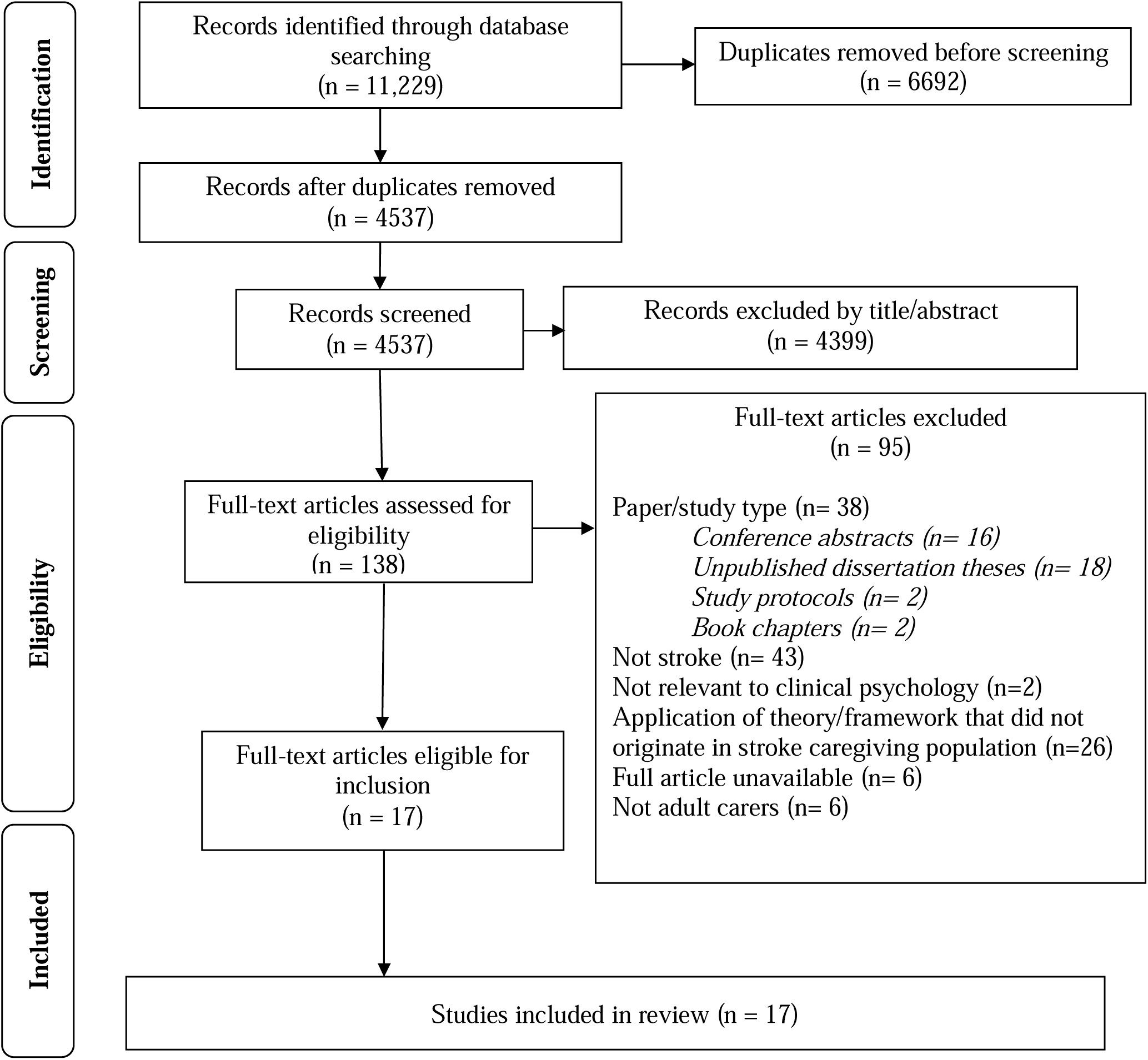
PRISMA diagram outlining identification of papers.

**Table 1:** Characteristics of the thirteen included theories, in chronological order of publication.

| No. | Theory Name | Topic/Focus | Study Characteristics |  |  |  |
| --- | --- | --- | --- | --- | --- | --- |
|  |  |  | Study author (year)<br>*Country* | Study Type | Population Demographics | Demographics |
| 1 | Maintaining caregiving at home | Experiences and perspectives of engaging in long-term in-home care provision | Subgranon & Lund (2000)<br>*Thailand* | Empirical | Family carers of stroke survivors | Thai adults |
| 2 | Model of quality of life for family caregivers of stroke survivors | Quality of life | White et al. (2004)<br>*United States* | Theoretical | Family/informal carers of stroke survivors | Unknown |
| 3 | Living with stroke - a theoretical framework of carers' experiences. | Carer needs and coping skills | Robinson et al. (2005)<br>*United Kingdom* | Theoretical | Family/informal carers of stroke survivors | Unknown |
| 4 | Conceptual model of influences of individual characteristics on quality-of-life (QoL) outcomes for stroke caregivers. | Stroke carer quality of life | Van Puymbroeck & Rittman (2005)<br>*United States* | Empirical | Informal carers of stroke survivors | Puerto Rican-Hispanic, African American, and Caucasian adults |
| 5 | Restructuring Life for Caregiving | Process of preparing for and beginning a new caregiving role | Silva-Smith (2007)<br>*United States* | Empirical | Family carers of stroke survivors | African American and Caucasian adults |
| 6 | “Timing It Right” (TIR) framework | Informal carers’ changing experiences and needs across the stroke trajectory | Cameron & Gignac (2008)<br>*Canada* | Theoretical | Literature on family carers of stroke survivors | Unknown/unspecified |
| 6a |  | Application of TIR framework | Cameron et al. (2013)<br>*Canada* | Empirical | Family/informal carers of stroke survivors | Unknown/unspecified |
| 6b |  |  | Yeung et al. (2015)<br>*Canada* | Empirical | Stroke survivors and their family carers | Chinese-Canadian adults |
| 6c |  |  | Burns et al. (2022)<br>*United States* | Empirical | Stroke survivors and their carers | African American adults |
| 6d |  |  | Shafer et al. (2023)<br>*United States* | Empirical | Care partners/ informal carers of people with aphasia after stroke | Adults of Black and White ethnicity |
| 7 | Seeking harmony to provide care for the stroke impaired | Coping strategies of carers | Lee & Mok (2011)<br>*China* | Empirical | Family carers of stroke impaired older relatives | Chinese adults |
| **8** | Stroke Crisis Trajectory | Challenges faced by carers following stroke | Lutz et al. (2011)<br>*United States* | Empirical | Stroke survivors and their family carers | Adults of Black, American Indian and White ethnicities |
| **9** | Model of the burden in caregivers of stroke survivors | Predictive factors of carer burden | Jaracz et al. (2012)<br>*Poland* | Empirical | Family carers of stroke survivors | Polish adults |
| **10** | Conceptual model of emotional vitality in informal caregivers | Emotional vitality in adopting carer role | Barbic et al. (2014)<br>*United States* | Empirical | Family/informal carers of stroke survivors | Unknown |
| **11** | Improving stroke caregiver readiness model | Improving stroke carer readiness for assuming caregiving role and identifying gaps in carer preparation | Lutz et al. (2017)<br>*United States* | Empirical | Family/informal carers of stroke survivors | Adults identifying with African American/Black, White or other ethnicities |
| **12** | Caregiver needs framework | Identifying carer needs to support engagement with support apps | Lobo et al. (2021)<br>*Australia/Denmark* | Theoretical | Family/informal carers of stroke survivors | Unknown |
| **13** | Experience trajectory of hospital-to-home transitional care of stroke survivors and family caregivers | Synthesis of experiences and difficulties during transition from hospital to home | Lin et al. (2022)<br>*China* | Empirical | Family carers of stroke survivors | Chinese adults |

The search strategy also returned papers presenting theories that were developed in other clinical populations (i.e. non-stroke populations) and had been subsequently applied to informal caregiving in stroke. To allow for a more meaningful focus to the review, and to ensure the validity and applicability of the synthesis, a retrospective decision was made for this review to focus on the seventeen papers that proposed and described theories that were developed in the target clinical population. Appendix 1 presents details of the papers featuring applied theories.

### 3.2 Thematic Synthesis

Seventeen papers met the inclusion criteria, and these presented thirteen distinct theories. Two overarching themes were developed (see Figure 3). The first, *Systemic and Cultural Factors*, appeared to shape the whole process and experience of informal caregiving. The second overarching theme, informal caregiving as *‘A staged process’,* contained four subthemes constituting stages across the inpatient and community settings that were relevant to carers: 1) *Stroke Event,* 2) *Discharge Preparation*, 3) *Initial Discharge Home,* and 4) *Long-term Home Living.* The two overarching themes contained three main themes: 1) *Adjustment to aspects of post-stroke life,* 2) *Emotional and psychological aspects of caregiving,* 3) *Carer Needs*.

**Figure 3:**
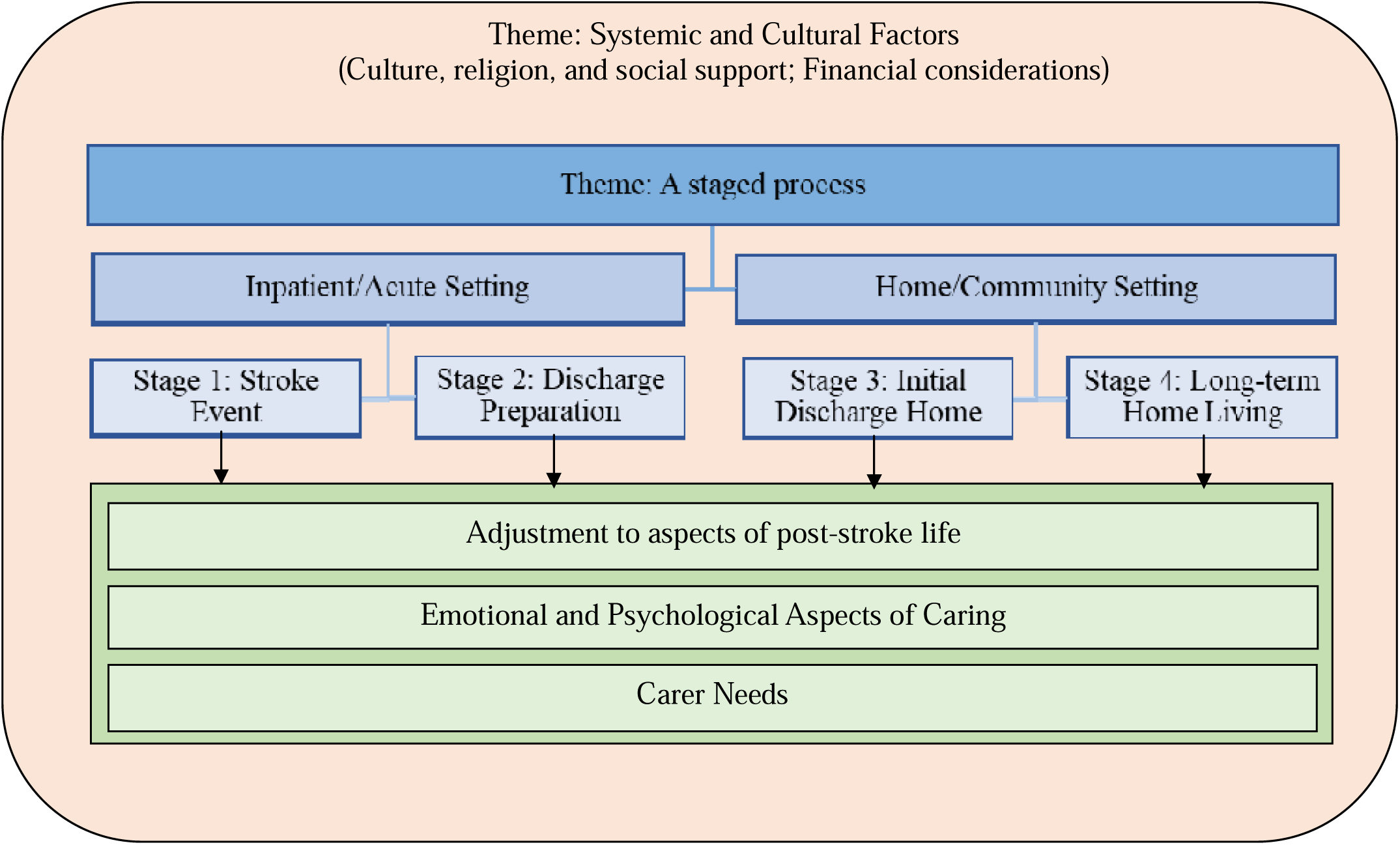
Diagram depicting themes developed through thematic synthesis.

Prior to exploring themes, the identified theories will be briefly summarised and compared. Four theories focussed on adapting to and maintaining the carer role (Subgranon & Lund, 2000; Lee & Mok, 2011; Jaracz et al., 2012; Barbic et al., 2014). Within these, theory 1 provided a broader focus (Subgranon & Lund, 2000) whereas theories 7, 9 and 10 focussed on the emotional and psychological components of role accommodation and maintenance (Lee & Mok, 2011; Jaracz et al., 2012; Barbic et al., 2014). Two theories, theory 2 (White et al., 2004) and theory 4 (Van Puymbroeck & Rittman, 2005), provided a focus on carer quality of life. The former conceptualised this from reviewing empirical studies focussing on the point of stroke event onwards (White et al., 2004). The latter quantitatively investigated predictive factors to form a theoretical explanatory quality of life model (Van Puymbroeck & Rittman, 2005). Theories 5 (Silva-Smith, 2007), 11 (Lutz et al., 2017) and 13 (Lin et al., 2022) broadly focussed on individuals’ preparedness to undertake the care role.

The dynamic and individual needs of carers were focussed on by theories 3, 6 and 12 (Robinson et al., 2005; Cameron & Gignac, 2008; Lobo et al., 2021). Theory 3 explored this through an empirical study of stroke carers, whereas theories 6 and 12 provided conceptualisations following reviews of empirical studies and relevant wider literature (Robinson et al., 2005; Cameron & Gignac, 2008; Lobo et al., 2021). Contrasting the other theories, theory 8 focussed on the challenging elements of adopting the carer role (Lutz et al., 2011). This concept was covered by the other theories yet was not the primary focus.

The two overarching themes developed from these theories will firstly be discussed. *Systemic and cultural* factors were core components of theories presented as shaping and influencing the whole experience of informal caregiving, by twelve papers. Theories highlighted the importance of understanding the role and influence of culture, faith, and religion in understanding the experience of caregiving after stroke (Burns et al., 2022; Lee & Mok, 2011; Subgranon & Lund, 2000). For example, certain cultures held expectations of family members to take on caregiving roles for other family members (Subgranon & Lund, 2000), which may influence their adjustment to the caregiving role. Religious beliefs and faith were identified as influential in approaches to caregiving roles and responsibilities, and to the emotional needs and wellbeing of carers (Burns et al., 2022). Family and social network support was indicated as important in many theories, influencing the carer’s ability to cope (Barbic et al., 2014), sense of isolation (Robinson et al., 2005) and overall quality of life (White et al., 2004). Many theories incorporated the importance of considering systemic factors such as financial implications of stroke and caregiving. Theories acknowledged the potential impact of these financial difficulties and considerations on the physical and mental health and wellbeing of carers (Lin et al., 2022; Lutz et al., 2011).

The second overarching theme, *A staged process*, contained four core stages across the stroke trajectory that helped to meaningfully organise the data. As shown in Figure 4, the included theories contributed data to these subthemes to differing extents. Some theories reflected the whole stroke trajectory, encompassing all four stages (e.g. White et al., 2004; Silva-Smith, 2007). Theories spanning two or more stages indicated carers may move through these stages differently, as opposed to them being prescriptive and linear. Other theories were narrower in scope, reflecting fewer subtheme stages (e.g. Subgranon & Lund, 2000; Van Puymbroeck & Rittman, 2005). The three main themes - *Adjustment to aspects of post-stroke life, Emotional and psychological aspects of caregiving, Carer Needs* - will now be discussed within the structure of these four stages.

**Figure 4:**
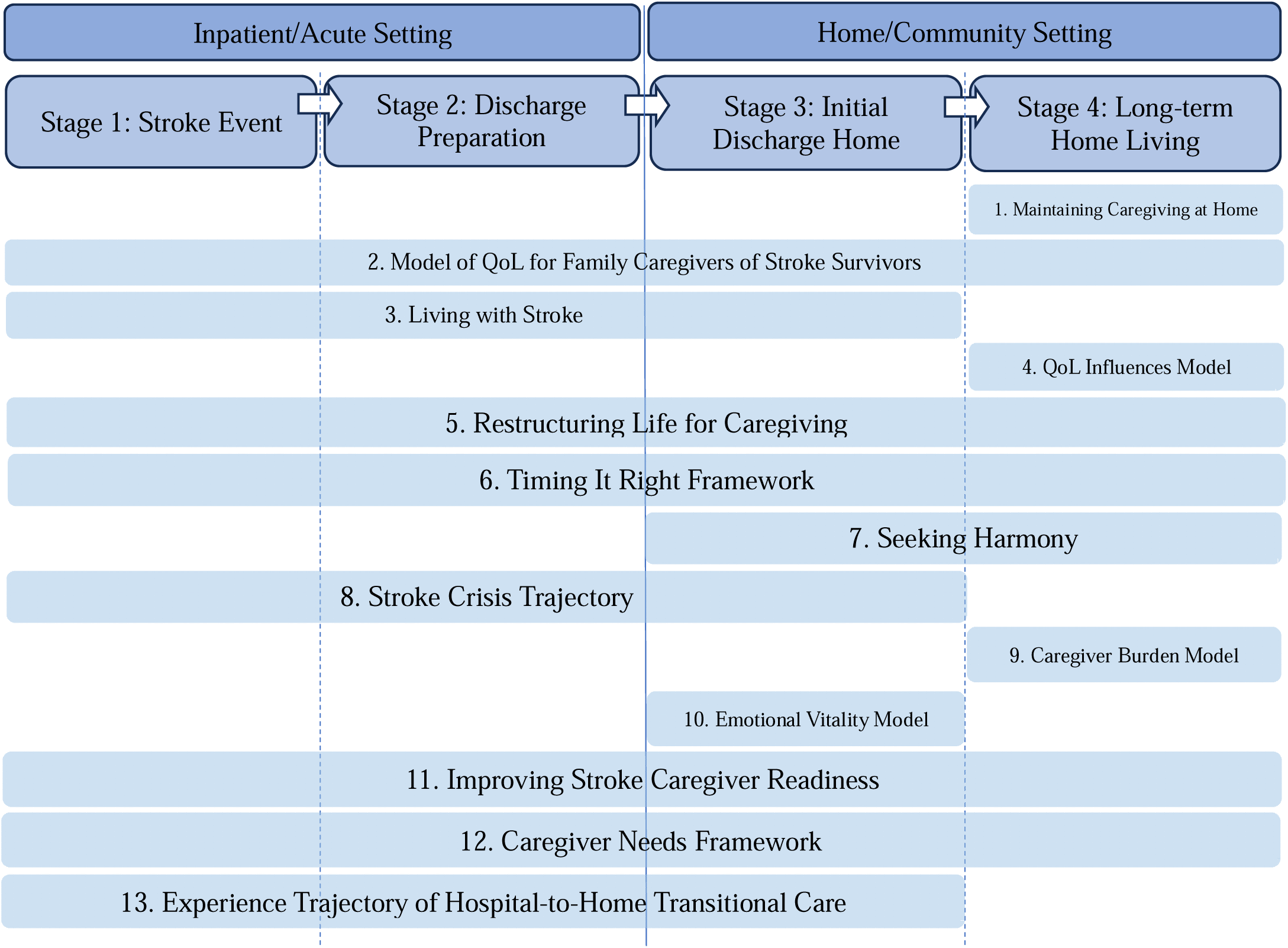
Visual depiction of stages covered by the thirteen theories included in the review. *Note: abbreviated theory titles have been used to promote figure readability*

Subtheme 1, *Stroke Event*, reflects the time surrounding stroke onset. Eleven theories contributed to the understanding of *Adjustment to aspects of post-stroke life* (Theme 1) at *Stage 1*. A common shared understanding between theories was that this first stage involves the carer processing and attempting to adjust to the initial shock and trauma caused by the stroke event (Robinson et al., 2005; Cameron & Gignac, 2008; Lutz et al., 2011; Lutz et al., 2017; Lin et al., 2022). Theories 6a-d also corroborate this understanding of *Adjustment to aspects of post-stroke life* at this stage. Adjusting to and processing the experience of stroke can also lead to carers having a present moment focus and limited consideration of the future (Subgranon & Lund, 2000; Lee & Mok, 2011). Theory 11 adds to this, proposing that any consideration of the future in this initial stage features unrealistic expectations for a return to pre-stroke life (Lutz et al., 2017). For theme 2, *Emotional and psychological aspects of caregiving*, seven theories contributed knowledge at *Stage 1*. Theories highlighted increased stress and anxiety for carers following the traumatic experience of stroke and an increased sense of uncertainty and unpredictability due to disrupted life plans and routine (Robinson et al., 2005; Cameron & Gignac, 2008; Lutz et al., 2011; Lin et al., 2022). Theory 13 also emphasised the likely conflicting emotions at this stage-continued shock and anxiety, alongside a sense of relief for the stroke survivor’s recovery (Lin et al., 2022). For theme 3, *Carer Needs*, five theories identified that in *Stage 1*, carers have informational needs pertaining to their loved one’s diagnosis and the impact of this on the stroke survivor and themselves (Silva-Smith, 2007; Cameron & Gignac, 2008; Cameron et al., 2013; Yeung et al., 2015; Burns et al., 2022; Lin et al., 2022). Two theories contributed the additional understanding of the importance of awareness of carers’ emotional needs to feel cared for and supported by others at this time (Burns et al., 2022; Lutz et al., 2011).

Subtheme 2, *Discharge Preparation*, is reflective of preparing for the stroke survivor’s discharge from inpatient into community or home-based settings. In relation to *Adjustment to aspects of post-stroke life*, theories propose that at this stage, carers are starting to develop an increased awareness of the impact of stroke on various elements of life, and the idea of reorganising life due to this (Silva-Smith, 2007; Cameron & Gignac, 2008; Lutz et al., 2017; Lin et al., 2022). Theory 3 specifies that this increased awareness includes adjusting to the idea of providing care for the stroke survivor (Robinson et al., 2005). Theory 13 suggests that during this stage, carers start to process the concept of the future being uncertain (Lin et al., 2022), yet other theories suggest this happens in the later stages (e.g. Cameron & Gignac, 2008; Lee & Mok, 2011). Considering theme 2, *Emotional and psychological aspects of caregiving*, several theories highlight that carers can experience varied and conflicting emotions at this stage, including hopes for the stroke survivor’s recovery simultaneous to anxiety at feeling unprepared for the future (Cameron & Gignac, 2008; Lutz et al., 2011; Lutz et al., 2017; Lin et al., 2022). Theory 3 also added that amidst this turbulence, carers often attempt to control their emotions to minimise any potential distress these may cause the stroke survivor (Robinson et al., 2005). *Carer Needs* during this stage were consistently reported to include training on care-related skills (e.g. Lutz et al., 2017; Lin et al., 2022). Several other theories also indicated that carers benefit from signposting to community support services to help them feel prepared for the return home (Cameron & Gignac, 2008; Lutz et al., 2011). Theory 11 added that carers also benefit from support to navigate and plan for the role changes they may experience (Lutz et al., 2017). Different from the other theories providing knowledge for this theme, theory 6b indicated that from *Stage 2* onwards, carers culture-specific needs become apparent, including translation or interpretation of information and support to appropriate languages and formats (Yeung et al., 2015).

Subtheme 3, *Initial Discharge Home*, refers to the stroke survivor’s initial return home or into a community setting. Theories collectively indicated that *Adjustment to aspects of post-stroke life* during *Stage 3* involved practising and applying care-related skills in the home environment (e.g. Robinson et al., 2005; Barbic et al., 2014). Carers also experience changes to their role and relationship with the stroke survivor and within the wider family system (e.g. Robinson et al., 2005; Silva-Smith, 2007). Return to the home environment also was theorised as resulting in carers realising how life may be different than they had expected, and adjusting to this loss of pre-stroke life (Robinson et al., 2005; Lutz et al., 2011; Lutz et al., 2017). *Emotional and psychological aspects of caregiving* at this stage were conceptualised to largely feature increased stress and anxiety as the extent of the care role becomes increasingly apparent (e.g. Robinson et al., 2005; Silva-Smith, 2007). Other theories added to this understanding that returning home from inpatient services can result in carers feeling isolated and alone (Robinson et al., 2005; Lutz et al., 2011). In relation to *Carer Needs,* theories covering this stage outlined the importance of carers recognising and meeting their own needs (Subgranon & Lund, 2000; Lutz et al., 2017), with them often needing support and encouragement to do this (Lee & Mok, 2011). Theories indicated that carers benefit from reassurance from healthcare professionals around their care-provision skills to enable them to maintain this role (Cameron & Gignac, 2008; Barbic et al., 2014). Social support from friends, family and the local community was consistently highlighted as beneficial to carers (e.g. Robinson et al., 2005; Lutz et al., 2011).

Subtheme 4, *Long-term Home Living*, reflects the period following initially taking on the care role, when the carer’s role becomes more established and longer term. Fewer theories contributed knowledge to this stage. Theories highlighted that at this stage, *Adjustment to aspects of post-stroke life* often features a stabilisation of the carer role and this becoming embedded within the carer’s life (Subgranon & Lund, 2000; Silva-Smith, 2007; Cameron & Gignac, 2008; Lee & Mok, 2011). This included carers developing an increased confidence in themselves and their abilities, in addition to working towards acceptance of the sense of future uncertainty they may experience (Cameron & Gignac, 2008; Lee & Mok, 2011). Focusses of theories relating to *Emotional and psychological aspects of caregiving* were that at this stage carers can often start to acknowledge some of the positive elements of their care role (Subgranon & Lund, 2000), and recognise the importance of meeting their own needs and wellbeing as their care role becomes more established (Cameron & Gignac, 2008; Lee & Mok, 2011). However, other theories noted that without the appropriate support, by this stage the effects of carer burden can become more apparent (Van Puymbroeck & Rittman, 2005; Jaracz et al., 2012). Important knowledge contributed by several theories relating to *Carer Needs,* was that some carers continue to struggle with recognising and prioritising their own needs in this stage and so often need support with this (Lee & Mok, 2011; Lutz et al., 2017). Connecting with other people in a similar situation through peer support was also noted as beneficial to carers, with emphasis placed on clinical services to help signpost to and facilitate this (Subgranon & Lund, 2000; Burns et al., 2022; Lee & Mok, 2011; Lutz et al., 2017).

Theories 2 and 12 contributed to our knowledge and understanding of the experience and psychological impact of providing informal care, yet provided a more generalised focus that didn’t differentiate between the stages (White et al., 2004; Lobo et al., 2021). The contributions of these theories will be briefly outlined. Both theories proposed that the *Adjustment to aspects of post-stroke life* for carers included their increased care-related roles and responsibilities (White et al., 2004; Lobo et al., 2021). As theory 12 offered a focus on carer needs, it added that part of the adjustment process includes their development of coping strategies to sustain the care role (Lobo et al., 2021). In relation to the *Emotional and psychological aspects of caregiving*, theory 2 indicated that carer quality of life is influenced by several care-related factors, including the extent of their care role and environmental factors such as social support and their relationship with the stroke survivor (Lobo et al., 2021). Theory 12’s contribution related to the emotional wellbeing of carers being promoted and maintained, by engagement with self-care and coping strategy development (Lobo et al., 2021). Considering *Carer Needs*, theory 2 emphasised the need for social support for carers from family and friends, in addition to carers’ informational needs relating to the impact and long-term management of stroke (White et al., 2004). Theory 12 corroborates this and added the need for carers needs to be managed (Lobo et al., 2021). Further information about the thematic synthesis findings can be seen in Table 2.

**Table 2:**
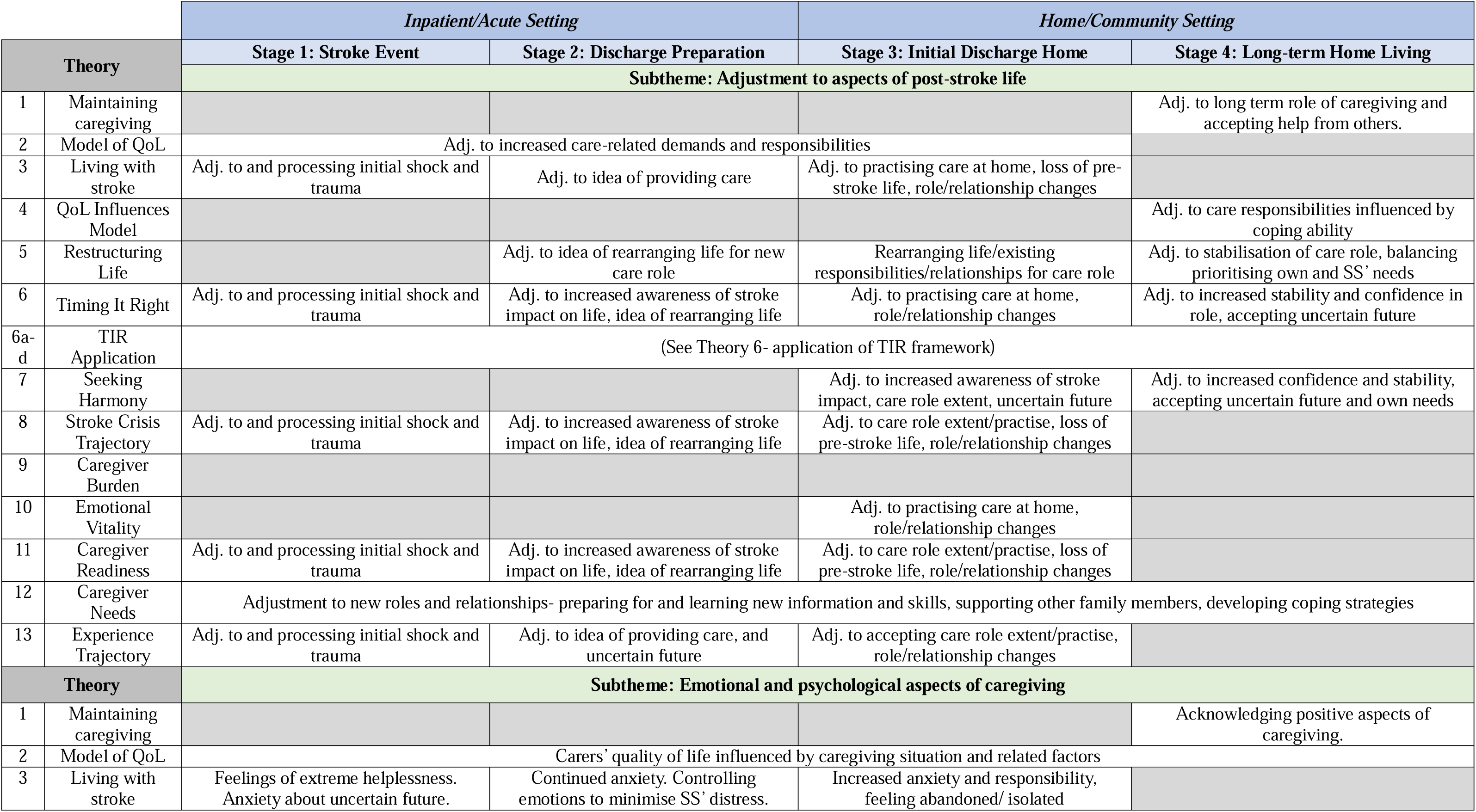
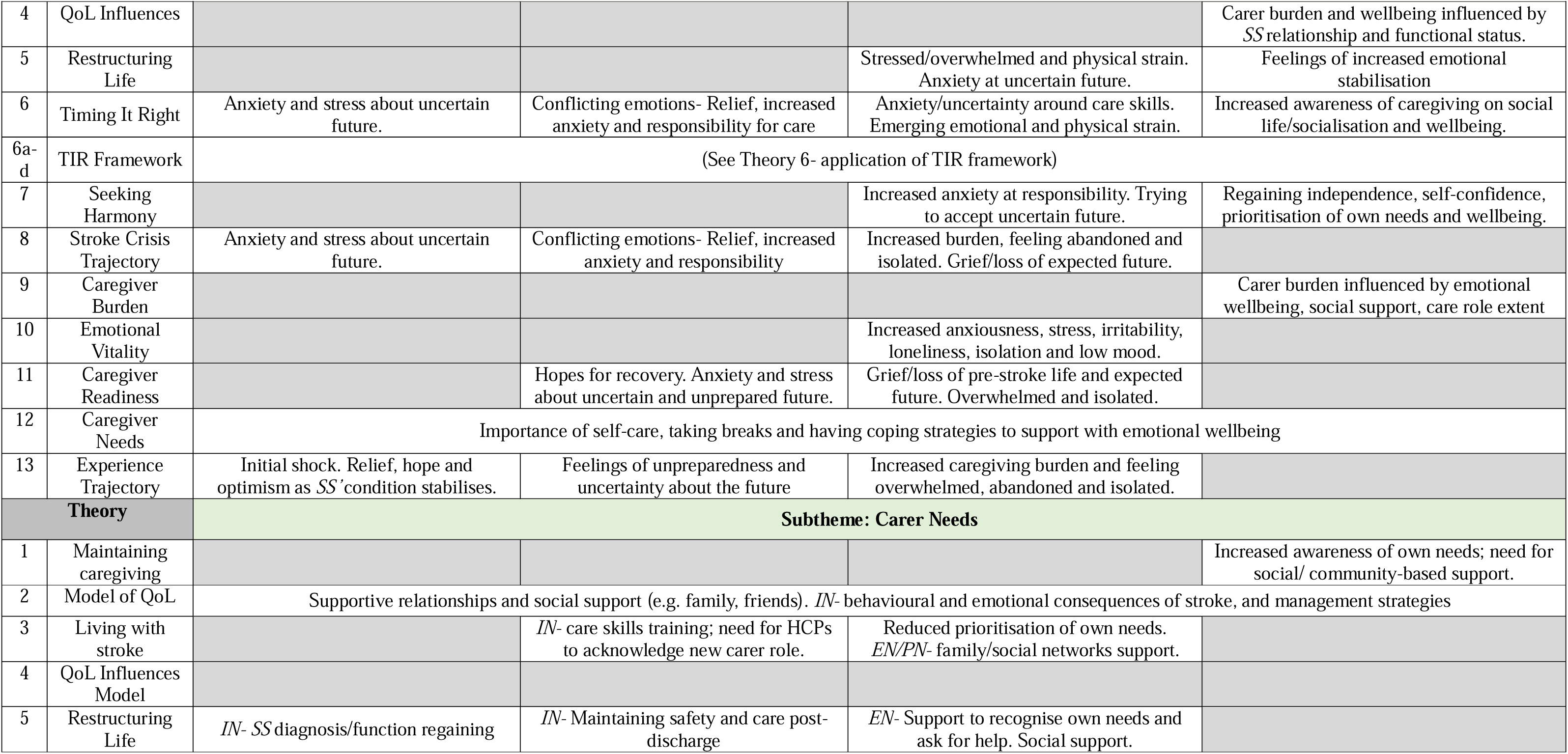
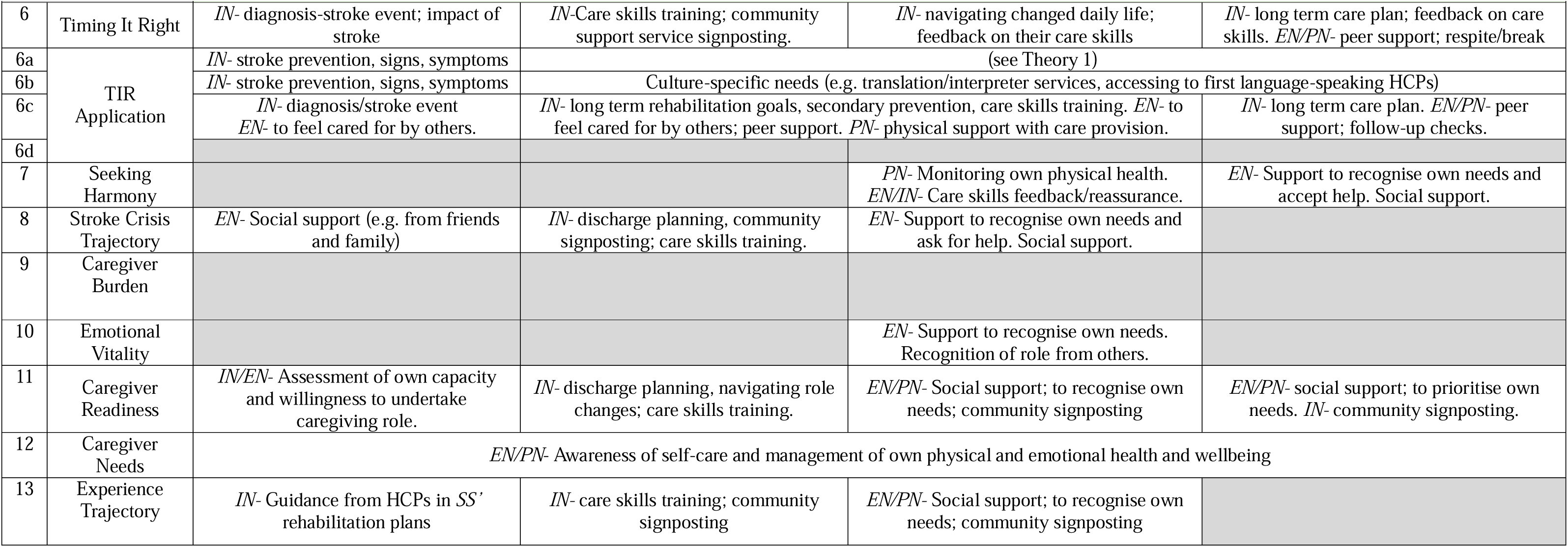
Thematic synthesis matrix of scoping review findings. *Note abbreviations: Adj.- adjustment; SS- stroke survivor; IN- informational/educational need; EN- emotional/psychological need; PN- physical need*

## 4. Discussion

This review discovered a significant number of theories existing in the current literature that relate to the experience and psychological impact of providing informal care in the neurological condition of stroke. Among the seventeen included papers, thirteen distinct theories were presented. Synthesising these theories resulted in the generation of new knowledge. To summarise this, the experience and psychological impact of informal caregiving was conceptualised as *A staged process*, comprising four main stages along the stroke trajectory of relevance to carers. *Systemic and cultural factors* (e.g. culture, faith and social support) shaped the experience and individuality of providing informal care. Overall, theories conceptualised the themes of *Adjustment to aspects of post-stroke life, Emotional and psychological aspects of caregiving,* and *Carer Needs* as being dynamic and evolving throughout the process of undertaking a caregiving role across the four stages.

Review findings will be discussed in relation to their positioning within and their contribution to the existing body of literature before discussion of their clinical implications.

This scoping review was the first of its kind, within the research team’s knowledge, to focus on the theories underpinning our understanding of the experience of informal care provision following stroke. This novel theoretical synthesis resulted in the conceptualisation of stroke informal caregiving as, *A staged process*, that closely mirrors the stages depicted in the literature as being experienced by the stroke survivor (Lutz et al., 2011). This indicates the relevance of parts of the stroke trajectory to the carer. Carers’ experiences and needs were found to be dynamic and change throughout these stages. This supports the proposition of LeLaurin et al. (2019) of the importance of theoretical knowledge that is reflective of the experience of informal caregiving specific to stroke as opposed to caregiving more generally.

Systemic and cultural factors were found to be influential to the caregiving experience. Drawing together multiple theories highlighted a commonality in different cultures having their own culture-specific needs and factors. For example, the cultural or religious beliefs of the carer and wider family system could shape their perception of the care-providing role and their responsibility to adopt this (Subgranon & Lund, 2000; Burns et al., 2022). This emphasises that to understand the individual carer experience following stroke, systemic factors relevant to their identity should be considered. Empirical studies on informal caregiving after stroke have similarly indicated that cultural values and beliefs are influential to motivations to undertaking the care role (Jullamate et al., 2007; Qiu et al., 2018). This is further reiterated in empirical literature on informal caregiving across health conditions (Zarzycki et al., 2023; Zarzycki et al., 2022). The present review therefore supports the existing literature indicating the importance of this concept, yet extend this by positioning it as a core component to theoretically understanding the caregiving experience and associated psychological impact. Relating to the importance of cultural considerations, this review is limited by only including English language papers. Papers proposing or describing theories from other countries and cultures may not have been identified. However, inclusion of multiple types and designs of study promoted review comprehensiveness.

The current review contributed to the wider literature a staged conceptualisation of the psychological adjustment process of carers. Adjustment as a psychological concept is relatively well understood in the literature. In line with existing psychological adjustment models to health conditions (Dekker & de Groot, 2018; Carroll et al., 2022; Taylor et al., 2011) adjustment is positioned in the present review as a dynamic process, shaped by a multitude of factors. Some models of adjustment after stroke exist, including Taylor et al.’s (2011) Social Cognitive Transition Model for Stroke, yet these relate to the adjustment process of a stroke survivor not a carer. Therefore, present review findings provide a specific focus on carer psychological adjustment. This means carer-specific experiences are identified, for example the learning and implementation of care-related skills (e.g. Cameron & Gignac, 2008; Barbic et al., 2014), that are not reflected in stroke survivor adjustment models (Taylor et al., 2011). The review synthesises theories to ascertain the role and experience of psychological adjustment for this population, and the stroke-specific factors (e.g. stroke trajectory stages) that are relevant to this process. Of relevance to clinical psychology, the psychological adjustment process by which an individual comes to see themselves as a ‘carer’, and how this change in their self-identity occurs, is not specifically covered within any of the identified theories and could comprise an area for future research.

The impact of caregiving and needs of stroke informal carers are well-evidenced within the literature (Ski & O’Connell, 2007; Zawawi et al., 2020). The present review therefore corroborates the importance of maintaining awareness of these factors and expands upon this by exploring and mapping how these needs change throughout the stroke trajectory, as proposed by the synthesis of multiple relevant theories. Whilst acknowledging the uniqueness and individuality of the caregiving experience, the review findings posit common emotional and psychological impacts and needs for informal stroke carers. It is clear that the psychological needs of carers are less extensively covered by current theories compared to other themes such as adjustment. This indicates the need for further research focussing on the psychological experiences and processes involved in adopting a carer role following stroke.

In summary, the review findings indicate that the themes and concepts of systemic and cultural factors, adjustment to post-stroke life, emotional and psychological aspects of caregiving, and the needs of informal carers of stroke survivors, are all important to the experience of providing informal care following stroke. However, theories adopt different points of focus and so address these themes to differing extents. A suggestion for future research is therefore for an overarching theory to be developed that effectively encompasses all these elements. The findings of the current review provide a starting point for the extension of this research.

### 4.1 Clinical implications

Review findings have several important implications for clinical practice. It has been demonstrated that carers’ psychological and emotional needs transition and change, corresponding with experiences along the stroke trajectory (e.g. returning home, long-term caregiving). Being aware of their changing needs and the factors impacting and shaping their experiences, can be helpful for clinicians in understanding the specific needs of this population.

To understand the individual experience of the carer in providing informal care following stroke, cultural and systemic factors influencing and shaping the carer’s identity and experience should be considered. Drawing together multiple theories highlighted a commonality in different cultures having their own culture-specific needs and factors. This is an important implication for clinical practice and services supporting carers.

Furthermore, holding an active awareness of cultural factors on the carer’s understanding, motivation for and experience of providing informal care may ensure culture-specific needs can be identified and addressed. At the service level, this awareness should be facilitated through staff training and reflective practice (Marshall et al., 2022). Historically there has been an under-representation of minority ethnic groups within research (Redwood, 2013). This, combined with the culture-specific needs identified as theoretically significant within the present review, further highlights the importance of including individuals from a diverse range of cultures and ethnicities within research. This is of core relevance within stroke research, due to ethnic minority groups being at higher risk of stroke (Fluck et al., 2023).

Although not included in the review, searches returned papers relating to stroke informal carers that applied theories from other disciplines and populations (Appendix 1). As multiple papers used these theories, the clinical relevance of these should be ascertained. Further research should explore the relevance of these theories to stroke carers, and any appropriate extensions or developments that could make them more reflective of this population.

A strength of the review is the enhancement of the scoping review methodology via inclusion of a second reviewer during all screening stages to reduce bias (Brett et al., 2014). However, little patient, carer, and public involvement (PCPI) may have limited this review, and inclusion of this may have enhanced the clinical applicability and impact of findings (Brett et al., 2014). If further research was to develop the overarching stroke caregiving theory, PCPI involvement should be a core part of this.

### 4.2 Conclusion

This scoping review identifies and synthesises theories of informal caregiving following stroke. Substantial theoretical knowledge exists within the current literature that can meaningfully contribute to the understanding of the experience and psychological impact of caregiving across the stroke pathway. The knowledge generated through this review can guide improvements in stroke services and clinical psychology practice. Further research should focus on the psychological needs and adjustment processes of stroke informal carers.

## 5. Declarations

### Author contributions

Bethany Harcourt conceptualised the review in collaboration with the wider research team, conducted the scoping review, thematic synthesis and drafted the first and subsequent manuscripts.

Audrey Bowen and Richard J Brown provided academic supervision throughout the project, provided guidance on the scoping review process, and contributed to the synthesis and interpretation of the data and revisions of drafts.

### Supplementary material

Supplementary material for this article is available in the appendices.

## Data Availability

All data produced in the present work are contained in the manuscript.

## Acknowledgements

The authors would like to thank Jade Wilkinson (JW) for her role of second reviewer during screening stages of the project, and both Michael Stevenson and Jessica Napthine-Hodgkinson, University of Manchester librarians, for their invaluable support and expertise in developing the search strategy used.

## Appendix 1 Table of papers returned by search strategy that applied a theory or framework to informal caregiving that did not originate from research in this population

*Note: these papers were not included in the review due to the theories and frameworks not being developed from research into informal caregiving in stroke. The table has been included here for interest of other researchers for future research*.

| No. | Theory/Framework Name | Topic/Focus | Study Characteristics |  |  |
| --- | --- | --- | --- | --- | --- |
|  |  |  | Study author (year)<br>*Country* | Population Demographics | Demographics |
| 1 | Cognitive adjustment theories | Psychological theories of adjusting to traumatic events | Thompson (1991)<br>*United States* | Informal caregivers of stroke survivors | Unknown/Not specified |
| 2 | Friedemann's framework of systemic organisation | Organising structure for nurses in meeting spiritual needs of patients | Pierce (2001)<br>*United States* | Family caregivers of stroke survivors | African American adults |
| 3 | Lazarus and Folkman's theory of stress and coping | Psychological theory of stress and coping style | Bakas & Burgener (2002) | Family caregivers of stroke survivors | Unknown/Not specified |
| 4 | Friedemann's framework of systemic organisation | Organising structure for nurses in meeting spiritual needs of patients | Pierce & Steiner (2004) | Family caregivers of stroke survivors | Unknown/Not specified |
| 5 | Friedemann's framework of systemic organisation | Organising structure for nurses in meeting spiritual needs of patients | Pierce et al. (2004)<br>*United States* | Family caregivers of stroke survivors | Unknown/Not specified |
| 6 | Orem's self-care deficit nursing theory | Self-care nursing theory | Cook et al. (2006)<br>*United States* | Family caregivers of stroke survivors | Adults identifying as American Indian, Alaska Native, Asian, Black or African American |
| 7 | Friedemann's framework of systemic organisation | Organising structure for nurses in meeting spiritual needs of patients | Pierce et al. (2007)<br>*United States* | Family caregivers of stroke survivors | Adults residing in United States |
| 8 | Friedemann's framework of systemic organisation | Organising structure for nurses in meeting spiritual needs of patients | Pierce et al. (2007)<br>*United States* | Family caregiver of stroke survivor | Caucasian male |
| **9** | Friedemann's framework of systemic organisation | Organising structure for nurses in meeting spiritual needs of patients | Pierce et al. (2008)<br>*Spain* | Family caregivers of stroke survivors | Adults of range of ethnicities (primarily Caucasian) |
| **10** | Orem's self-care deficit nursing theory | Self-care nursing theory | Steiner et al. (2008)<br>*United States* | Family caregivers of stroke survivors | Adults from a range of ethnicities |
| **11** | Friedemann's framework of systemic organisation | Organising structure for nurses in meeting spiritual needs of patients | Pierce et al. (2010)<br>*United States* | Family caregivers of stroke survivors | Unknown/Not specified |
| **12** | Supportive Care Needs Framework | Conceptualisation of domains impacting on patients' and support persons' needs | MacIsaac et al. (2011)<br>*Canada* | Family caregivers of stroke survivors | Unknown/Not specified |
| **13** | Social Exchange Theory | Psychological theory of social relationships | Sakunhongsoophon et al. (2011)<br>*Thailand* | Family caregivers of stroke survivors | Thai adults |
| **14** | Lazarus and Folkman's theory of stress and coping | Psychological theory of stress and coping style | Gonzalez & Bakas (2013)<br>*United States* | Family caregivers of stroke survivors | Adults identifying as African American, Caucasian and Asian |
| **15** | Schaefer and Moos, and Tedeschi and Calhoun post-traumatic growth models | Psychological theory originating from trauma research | Hallam & Morris (2014)<br>*United Kingdom* | Informal caregivers of stroke survivors | Unknown/Not specified |
| **16** | Theory of humanising care | Theoretical nursing framework for delivering care | Pound & Greenwood (2016)<br>*United Kingdom* | Individuals providing informal care for someone after a stroke | Black and minority ethnic groups |
| **17** | Friedemann's framework of systemic organisation | Organising structure for nurses in meeting spiritual needs of patients | Pierce et al. (2019)<br>*United States* | Male family caregivers of stroke survivors | Unknown/Not specified |
| **18** | Theory of social representations | Social psychological approach for construction of meaning | Dos Santos Alves et al. (2021)<br>*Brazil* | Stroke patients' family caregivers | Adults from Brazil |
| **19** | Intersectional framework | Systemic framework incorporating concepts such as socioeconomic, emotional and health factors | Moore et al. (2021)<br>*United States/Ghana* | Family caregivers of stroke survivors | African adults residing in Lomé, Togo |
| **20** | Theory of unitary caring | Philosophical framework for healthcare practice | Alselami & Butcher (2022)<br>*United States/Saudi Arabia* | Family caregivers of patients hospitalised with stroke in Saudi Arabia | Adults from Saudi Arabia |
| **21** | Lazarus and Folkman's theory of stress and coping | Psychological theory of stress and coping style | Lee & Song (2022)<br>*South Korea* | Informal caregivers of stroke survivors | Unknown/Not specified |

## References

Achilike, S., Beauchamp, J. E., Cron, S. G., Okpala, M., Payen, S. S., Baldridge, L., Okpala, N., Montiel, T. C., Varughese, T., Love, M., Fagundes, C., Savitz, S., & Sharrief, A. (2020). Caregiver burden and associated factors among informal caregivers of stroke survivors. Journal of Neuroscience Nursing, 52(6), 277–283. DOI: 10.1097/JNN.0000000000000552

Aranda, M. P., & Knight, B. G. (1997). The influence of ethnicity and culture on the caregiver stress and coping process: a sociocultural review and analysis. The Gerontologist, 37(3), 342–354. 10.1093/geront/37.3.342

Alselami, S., & Butcher, H. K. (2022). A Unitary Caring Theory Perspective of Family Caregiving for Patients Hospitalized With a Stroke in Saudi Arabia. Nursing Science Quarterly, 35(2), 191–202. 10.1177/08943184211070608

Bakas, T., & Burgener, S. C. (2002). Predictors of emotional distress, general health, and caregiving outcomes in family caregivers of stroke survivors. Topics in Stroke Rehabilitation, 9(1), 34–45. 10.1310/GN0J-EXVX-KX0B-8X43

Barbic, S. P., Mayo, N. E., White, C. L., & Bartlett, S. J. (2014). Emotional vitality in family caregivers: content validation of a theoretical framework. Quality of Life Research, 23, 2865–2872.

Barnett-Page, E., & Thomas, J. (2009). Methods for the synthesis of qualitative research: a critical review. BMC medical research methodology, 9(1), 1–11. 10.1186/1471-2288-9-59

Belur, J., Tompson, L., Thornton, A., & Simon, M. (2021). Interrater reliability in systematic review methodology: exploring variation in coder decision-making. Sociological Methods & Research, 50(2), 837–865. 10.1177/0049124118799372

Bray, B. D., Paley, L., Hoffman, A., James, M., Gompertz, P., Wolfe, C. D., Hemingway, H., & Rudd, A. G. (2018). Socioeconomic disparities in first stroke incidence, quality of care, and survival: a nationwide registry-based cohort study of 44 million adults in England. The Lancet Public Health, 3(4), e185–e193. 10.1016/S2468-2667(18)30030-6

Brereton, L., Carroll, C., & Barnston, S. (2007). Interventions for adult family carers of people who have had a stroke: a systematic review. Clinical Rehabilitation, 21(10), 867–884. 10.1177/0269215507078313

Brett, J. O., Staniszewska, S., Mockford, C., HerronLJMarx, S., Hughes, J., Tysall, C., & Suleman, R. (2014). Mapping the impact of patient and public involvement on health and social care research: a systematic review. Health Expectations, 17(5), 637–650. 10.1111/j.1369-7625.2012.00795.x

British Psychological Society. (2017). Practice Guidelines-Third Edition. https://explore.bps.org.uk/content/report-guideline/bpsrep.2017.inf115

British Psychological Society. (2023). Recommendations for Integrated Community Stroke Services: Service design, workforce planning & clinical governance requirements for a high-quality service and rehabilitation outcomes. https://cms.bps.org.uk/sites/default/files/2023-04/BRE56%20Recommendations%20for%20Integrated%20Community%20Stroke%20Services_April.pdf

Burns, S. P., Lutz, B. J., & Magwood, G. S. (2022). ‘Timing it Right’: needs of African American adults with stroke and their caregivers across the care continuum. Ethnicity & Health, 27(2), 420–434. 10.1080/13557858.2019.1693512

Campbell, M., Fitzpatrick, R., Haines, A., Kinmonth, A. L., Sandercock, P., Spiegelhalter, D., & Tyrer, P. (2000). Framework for design and evaluation of complex interventions to improve health. Bmj, 321(7262), 694–696. 10.1136/bmj.321.7262.694

Cameron, J. I., & Gignac, M. A. (2008). “Timing It Right”: A conceptual framework for addressing the support needs of family caregivers to stroke survivors from the hospital to the home. Patient Education and Counseling, 70(3), 305–314. 10.1016/j.pec.2007.10.020

Cameron, J. I., Naglie, G., Silver, F. L., & Gignac, M. A. (2013). Stroke family caregivers’ support needs change across the care continuum: a qualitative study using the timing it right framework. Disability and Rehabilitation, 35(4), 315–324. 10.3109/09638288.2012.691937

Carroll, S., Moon, Z., Hudson, J., Hulme, K., & Moss-Morris, R. (2022). An Evidence-Based Theory of Psychological Adjustment to Long-Term Physical Health Conditions: Applications in Clinical Practice. Psychosomatic Medicine, 84(5), 547–559. 10.1097/PSY.0000000000001076

Clarke, D. J., Hawkins, R., Sadler, E., Harding, G., McKevitt, C., Godfrey, M., Dickerson, J., Farrin, A. J., Kalra, L., Smithard, D., & Forster, A. (2014). Introducing structured caregiver training in stroke care: findings from the TRACS process evaluation study. BMJ open, 4(4), e004473. doi:10.1136/bmjopen-2013-004473

Craig, P., Dieppe, P., Macintyre, S., Michie, S., Nazareth, I., & Petticrew, M. (2008). Developing and evaluating complex interventions: the new Medical Research Council guidance. BMJ, 337. 10.1136/bmj.a1655

Cook, A. M., Pierce, L. L., Hicks, B., & Steiner, V. (2006). Self-care needs of caregivers dealing with stroke. Journal of Neuroscience Nursing, 38(1), 31–36. 10.1097/01376517-200602000-00007

Creasy, K. R., Lutz, B. J., Young, M. E., & Stacciarini, J. M. R. (2015). Clinical implications of familyLJcentered care in stroke rehabilitation. Rehabilitation Nursing, 40(6), 349–359. 10.1002/rnj.188

Davies, P., Walker, A. E., & Grimshaw, J. M. (2010). A systematic review of the use of theory in the design of guideline dissemination and implementation strategies and interpretation of the results of rigorous evaluations. Implementation science, 5, 1–6. 10.1186/1748-5908-5-14

Dekker, J., & de Groot, V. (2018). Psychological adjustment to chronic disease and rehabilitation–an exploration. Disability and Rehabilitation, 40(1), 116–120. 10.1080/09638288.2016.1247469

Denham, A. M., Wynne, O., Baker, A. L., Spratt, N. J., Loh, M., Turner, A., Magin, P., & Bonevski, B. (2022). The long-term unmet needs of informal carers of stroke survivors at home: a systematic review of qualitative and quantitative studies. Disability and rehabilitation, 44(1), 1–12. 10.1080/09638288.2020.1756470

dos Santos Alves, P., Dias da Silva, S. É., Santos Araújo, J., Farias da Cunha, N. M., Alves Mour, A. A., & Lobato da Costa, J. (2021). Caring for Oneself: Social Representations of Stroke Patients’ Family Caregivers. Revista de Pesquisa: Cuidado e Fundamental, 13(1).

Eldred, C., & Sykes, C. (2008). Psychosocial interventions for carers of survivors of stroke: a systematic review of interventions based on psychological principles and theoretical frameworks. British journal of health psychology, 13(3), 563–581. 10.1348/135910707x236899

Elvish, R., Lever, S. J., Johnstone, J., Cawley, R., & Keady, J. (2013). Psychological interventions for carers of people with dementia: A systematic review of quantitative and qualitative evidence. Counselling and Psychotherapy Research, 13(2), 106–125.

Felix, M. S., Le, T. N. P., Wei, M., & Puspitasari, D. C. (2021). Scoping review: Health needs of the family caregivers of elderly stroke survivors. Health & Social Care in the Community, 29(6), 1683–1694. 10.1111/hsc.13371

Feng, G.C. Factors affecting intercoder reliability: a Monte Carlo experiment. Qual Quant 47, 2959–2982 (2013). 10.1007/s11135-012-9745-9

Fluck, D., Fry, C. H., Gulli, G., Affley, B., Robin, J., Kakar, P., … & Han, T. S. (2023). Adverse stroke outcomes amongst UK ethnic minorities: a multi-centre registry-based cohort study of acute stroke. Neurological Sciences, 1–10. 10.1007/s10072-023-06640-z

Ghazzawi, A., Kuziemsky, C., & O’Sullivan, T. (2016). Using a complex adaptive system lens to understand family caregiving experiences navigating the stroke rehabilitation system. BMC Health Services Research, 16, 1–10. 10.1186/s12913-016-1795-6

Gittins, M., Lugo-Palacios, D., Vail, A., Bowen, A., Paley, L., Bray, B., & Tyson, S. (2021). Stroke impairment categories: A new way to classify the effects of stroke based on stroke-related impairments. Clinical Rehabilitation, 35(3), 446–458. 10.1177/0269215520966473

Gonzalez, C., & Bakas, T. (2013). Factors associated with stroke survivor behaviors as identified by family caregivers. Rehabilitation Nursing, 38(4), 202–211. 10.1002/rnj.85

Greenwood, N., Mackenzie, A., Cloud, G. C., & Wilson, N. (2008). Informal carers of stroke survivors–factors influencing carers: a systematic review of quantitative studies. Disability and Rehabilitation, 30(18), 1329–1349. 10.1080/09638280701602178

Hallam, W., & Morris, R. (2014). PostLJtraumatic growth in stroke carers: A comparison of theories. British Journal of Health Psychology, 19(3), 619–635. 10.1111/bjhp.12064

Harrison, M., & Palmer, R. (2015). Exploring patient and public involvement in stroke research: a qualitative study. Disability and Rehabilitation, 37(23), 2174–2183.

Hussain, N. A., Abdullah, M. R., Esa, A. R., Mustapha, M., & Yusoff, N. (2014). Predictors of life satisfaction among family caregivers of hospitalized first-ever stroke patients in Kelantan. Caring, 18, 19. 10.3109/09638288.2014.1001525

Intercollegiate Stroke Working Party. (2012). National clinical guideline for stroke. http://www.askdoris.org/guidelines/RCP/National-Clinical-Guidelines-for-Stroke-Fourth-Edition.pdf

Jaracz, K., Grabowska-Fudala, B., & Kozubski, W. (2012). Caregiver burden after stroke: towards a structural model. Neurologia i neurochirurgia polska, 46(3), 224–232. 10.5114/ninp.2012.29130

Jo□nsson, A. C., Lindgren, I., Hallstro□m, B., Norrving, B., & Lindgren, A. (2005). Determinants of quality of life in stroke survivors and their informal caregivers. Stroke, 36(4), 803–808. 10.1161/01.str.0000160873.32791.20

Jullamate, P., de Azeredo, Z., Rosenberg, E., Pàul, C., & Subgranon, R. (2007). Informal stroke rehabilitation: what are the main reasons of Thai caregivers?. International Journal of Rehabilitation Research, 30(4), 315–320. 10.1097/mrr.0b013e3282f14539

Keaton, L., Pierce, L. L., Steiner, V., Lance, K., Masterson, M., Rice, M. S., & Smith, J. L. (2004). An E-rehabilitation team helps caregivers deal with stroke. Internet Journal of Allied Health Sciences and Practice, 2(4), 7. DOI: 10.46743/1540-580X/2004.1057

Kerr, S. M., & Smith, L. N. (2001). Stroke: an exploration of the experience of informal caregiving. Clinical rehabilitation, 15(4), 428–436. 10.1191/026921501678310234

Krieger, T., Jungbauer, J., Feron, F., & Dorant, E. (2015). Developing a Complex Intervention Program for Informal Caregivers of Stroke Survivors: Theresia Krieger. The European Journal of Public Health, 25(suppl_3), ckv175-225. 10.1093/eurpub/ckv175.225

Krishnan, S., Pappadis, M. R., Weller, S. C., Stearnes, M., Kumar, A., Ottenbacher, K. J., & Reistetter, T. A. (2017). Needs of stroke survivors as perceived by their caregivers: a scoping review. American Journal of Physical Medicine & Rehabilitation, 96(7), 487. 10.1097/phm.0000000000000717

Lazarus, R. S., and Folkman, S. (1984). Stress, Appraisal, and Coping. New York: Springer.

Leamy, M., Bird, V., Le Boutillier, C., Williams, J., & Slade, M. (2011). Conceptual framework for personal recovery in mental health: systematic review and narrative synthesis. The British Journal of Psychiatry : the Journal of Mental Science, 199(6), 445–452. 10.1192/bjp.bp.110.083733

Lee, R. L., & Mok, E. S. (2011). Seeking harmony in the provision of care to the strokeLJimpaired: views of Chinese family caregivers. Journal of Clinical Nursing, 20(9LJ10), 1436–1444. 10.1111/j.1365-2702.2010.03500.x

Lee, Y., & Song, Y. (2022). Coping as a Mediator of the Relationship between Stress and Anxiety in Caregivers of Patients with Acute Stroke. Clinical Nursing Research, 31(1), 136–143. 10.1177/10547738211021223

LeLaurin, J., Schmitzberger, M., Eliazar-Macke, N., Freytes, I. M., Dang, S., & Uphold, C. (2019). A commentary on methodological issues in stroke caregiver research: lessons learned from three RESCUE intervention studies. Topics in Stroke Rehabilitation, 26(5), 399–404. 10.1080/10749357.2019.1607485

Levac, D., Colquhoun, H., & O’Brien, K. K. (2010). Scoping studies: advancing the methodology. Implementation Science, 5, 1–9. 10.1186/1748-5908-5-69

Lin, S., Wang, C., Wang, Q., Xie, S., Tu, Q., Zhang, H., … & Redfern, J. (2022). The experience of stroke survivors and caregivers during hospital-to-home transitional care: A qualitative longitudinal study. International Journal of Nursing Studies, 130, 104213. 10.1016/j.ijnurstu.2022.104213

Lobo, E. H., Frølich, A., Kensing, F., Rasmussen, L. J., Livingston, P. M., Grundy, J., & Abdelrazek, M. (2021). mHealth applications to support caregiver needs and engagement during stroke recovery: a content review. Research in Nursing & Health, 44(1), 213–225. 10.1002/nur.22096

Lutz, B. J., Young, M. E., Cox, K. J., Martz, C., & Creasy, K. R. (2011). The crisis of stroke: experiences of patients and their family caregivers. Topics in Stroke Rehabilitation, 18(6). 10.1310/tsr1806-786

Lutz, B. J., Young, M. E., Creasy, K. R., Martz, C., Eisenbrandt, L., Brunny, J. N., & Cook, C. (2017). Improving stroke caregiver readiness for transition from inpatient rehabilitation to home. The Gerontologist, 57(5), 880–889. 10.1093/geront/gnw135

MacIsaac, L., Harrison, M. B., Buchanan, D., & Hopman, W. M. (2011). Supportive care needs after an acute stroke: a descriptive enquiry of caregivers’ perspective. Journal of Neuroscience Nursing, 43(3), 132–140. 10.1097/jnn.0b013e3182135b28

Marshall, T., Keville, S., Cain, A., & Adler, J. R. (2022). Facilitating reflection: A review and synthesis of the factors enabling effective facilitation of reflective practice. Reflective Practice, 23(4), 483–496. 10.1080/14623943.2022.2064444

Michie, S., & Prestwich, A. (2010). Are interventions theory-based? Development of a theory coding scheme. Health psychology, 29(1), 1. 10.1037/a0016939

Moore, A. R., Yaa Owusu, A., Moore, S., & Knight, R. (2021). Caring for a Loved One with Stroke in Lomé, Togo: an Intersectional Framework. Ageing International, 1–15. 10.1007/s12126-021-09427-9

Moore, G. F., Audrey, S., Barker, M., Bond, L., Bonell, C., Hardeman, W., Moore, L., O’Cathain, A., Tinati, T., Wight, D., & Baird, J. (2015). Process evaluation of complex interventions: Medical Research Council guidance. BMJ, 350. 10.1136/bmj.h1258

Moura, A., Teixeira, F., Amorim, M., Henriques, A., Nogueira, C., & Alves, E. (2022). A scoping review on studies about the quality of life of informal caregivers of stroke survivors. Quality of Life Research, 1–20. 10.1007/s11136-021-02988-x

Muhrodji, P., Wicaksono, H. D. A., Satiti, S., Trisnantoro, L., Setyopranoto, I., & Vidyanti, A. N. (2021). Roles and problems of stroke caregivers: a qualitative study in Yogyakarta, Indonesia. F1000Research, 10. 10.12688/f1000research.52135.2

National Institute for Health and Care Excellence. (2020). Supporting adult carers. [NICE Guideline No. 150]. https://www.nice.org.uk/guidance/NG150

National Institute for Health and Care Excellence. (2023). Stroke rehabilitation in adults. [NICE Guideline No. 236]. https://www.nice.org.uk/guidance/ng236

NHS England and NHS Improvement. (2021). National stroke service model: integrated stroke delivery networks. London: National Stroke Programme NHS England and NHS Improvement. https://www.england.nhs.uk/publication/national-stroke-service-model-integrated-stroke-delivery-networks/

Orwin R. G. (1994). Evaluating coding decisions. In Cooper H., Hedges L. V. (Eds.), The handbook of research synthesis (pp. 139–162). New York, NY: Russell Sage.

Peters MDJ, Godfrey C, McInerney P, Munn Z, Tricco AC, Khalil, H. Chapter 11: Scoping Reviews (2020 version). In: Aromataris E, Munn Z (Editors). JBI Manual for Evidence Synthesis, JBI, 2020. Available from https://synthesismanual.jbi.global. 10.46658/JBIMES-20-12

Peters, M. D., Marnie, C., Colquhoun, H., Garritty, C. M., Hempel, S., Horsley, T., Langlois, E. V., Lillie, E., O’Brien, K. K., Tunçalp, Ö., Wilson, M. G.,, Zarin, W., & Tricco, A. C. (2021). Scoping reviews: reinforcing and advancing the methodology and application. Systematic Reviews, 10(1), 1–6. 10.1186/s13643-021-01821-3

Pierce, L. L., Steiner, V., Alamina, F., Onyekelu, D., & Stevenson, S. (2019). Male caregivers report problems in caring at home after spouses survive stroke. Home healthcare now, 37(1), 23–32. 10.1097/nhh.0000000000000705

Pierce, L. L., Steiner, V., Govoni, A., Thompson, T. C., & Friedemann, M. L. (2007). Two sides to the caregiving story. Topics in Stroke Rehabilitation, 14(2), 13–20. 10.1310/tsr1402-13

Pierce, L. L., Steiner, V., Govoni, A. L., Hicks, B., Thompson, T. L. C., & Friedemann, M. L. (2004). Caregivers dealing with stroke pull together and feel connected. Journal of Neuroscience Nursing, 36(1), 32–39. 10.1097/01376517-200402000-00005

Pierce, L. L. (2001). Caring and expressions of spirituality by urban caregivers of people with stroke in African American families. Qualitative Health Research, 11(3), 339–352. 10.1177/104973230101100305

Pierce, L. L., & Steiner, V. (2004). What are male caregivers talking about?. Topics in Stroke Rehabilitation, 11(2), 77–83. 10.1310/mhrl-nwl5-9xcg-jemc

Pierce, L. L., Steiner, V., Seymour, J. R., Wicks, B., Wright, C., & Thompson, T. (2010). Questions caregivers asked in caring for persons with stroke. Online Journal of Nursing Informatics, 14(2), 1–20.

Pierce, L. L., Steiner, V., Havens, H., & Tormoehlen, K. (2008). Spirituality expressed by caregivers of stroke survivors. Western Journal of Nursing Research, 30(5), 606–619. 10.1177/0193945907310560

Pierce, L. L., Thompson, T. L., Govoni, A. L., & Steiner, V. (2012). Caregivers’ incongruence: emotional strain in caring for persons with stroke. Rehabilitation Nursing, 37(5), 258–266. 10.1002/rnj.35

Pierce, L. L., Steiner, V., Hicks, B., & Dawson-Weiss, J. (2007). Perceived experience of caring for a wife with stroke. Rehabilitation Nursing Journal, 32(1), 35–40. 10.1002/j.2048-7940.2007.tb00147.x

Pierce, L. L., Steiner, V., Govoni, A. L., Hicks, B., Thompson, T. L. C., & Friedemann, M. L. (2004). Internet-based support for rural caregivers of persons with stroke shows promise. Rehabilitation Nursing Journal, 29(3), 95–99. 10.1002/j.2048-7940.2004.tb00319.x

Pierce, L. L. (2001). Caring and expressions of stability by urban family caregivers of persons with stroke within African American family systems. Rehabilitation Nursing, 26(3), 100–116. 10.1002/j.2048-7940.2001.tb02213.x

Pierce, L. L., Steiner, V., Cervantez Thompson, T. L., & Friedemann, M. L. (2014). Linking theory with qualitative research through study of stroke caregiving families. Rehabilitation Nursing, 39(3), 157–165. 10.1002/rnj.83

Pomerantz, A. M. (2016). Clinical psychology: Science, practice, and culture. Sage Publications.

Pound, C., & Greenwood, N. (2016). The human dimensions of post-stroke homecare: experiences of older carers from diverse ethnic groups. Disability and Rehabilitation, 38(20), 1987–1999. 10.3109/09638288.2015.1107783

Qiu, X., Sit, J. W., & Koo, F. K. (2018). The influence of Chinese culture on family caregivers of stroke survivors: a qualitative study. Journal of Clinical Nursing, 27(1-2), e309–e319. 10.1111/jocn.13947

QSR International (2018). NVIVO, QSR International Pty Ltd. Version 12. Retrieved from: 60 https://www.qsrinternational.com/nvivo/home.

Redfern, J., McKevitt, C., & Wolfe, C. D. (2006). Development of complex interventions in stroke care: a systematic review. Stroke, 37(9), 2410–2419. DOI: 10.1161/01.STR.0000237097.00342.a9

Robinson, L., Francis, J., James, P., Tindle, N., Greenwell, K., & Rodgers, H. (2005). Caring for carers of people with stroke: developing a complex intervention following the Medical Research Council framework. Clinical Rehabilitation, 19(5), 560–571. 10.1191/0269215505cr787oa

Sakunhongsophon, S., Sirapo-ngam, Y., Tripp-Reimer, T., & Junda, T. (2011). Stroke caregiving networks in Bangkok: patterns of social exchange behavior. Pacific Rim International Journal of Nursing Research, 15(2), 152–165.

Sentinel Stroke National Audit Programme (SSNAP) Annual Report 2023. London: King’s College London, 2023. https://www.strokeaudit.org/About/About-SSNAP/Annual-reports.aspx

Shafer, J. S., Haley, K. L., & Jacks, A. (2023). Barriers to Informational Support for Care Partners of People With Aphasia After Stroke. American Journal of Speech-Language Pathology, 32(5), 2211–2231. 10.1044/2023_ajslp-22-00391

Sheldon, K., & Harding, E. (2010). Good practice guidelines to support the involvement of service users and carers in clinical psychology services. Leicester, UK: British Psychological Society

Silva-Smith, A. L. (2007). Restructuring life: Preparing for and beginning a new caregiving role. Journal of family nursing, 13(1), 99–116. 10.1177/1074840706297425

Ski, C., & O’Connell, B. (2007). Stroke: the increasing complexity of carer needs. Journal of Neuroscience Nursing, 39(3), 172–179.

Smeeton, N. C., Heuschmann, P. U., Rudd, A. G., McEvoy, A. W., Kitchen, N. D., Sarker, S. J., & Wolfe, C. D. (2007). Incidence of hemorrhagic stroke in black Caribbean, black African, and white populations: the South London stroke register, 1995-2004. Stroke, 38(12), 3133–3138. 10.1161/STROKEAHA.107.487082

Spring, B. (2007). EvidenceLJbased practice in clinical psychology: What it is, why it matters; what you need to know. Journal of clinical psychology, 63(7), 611–631. 10.1002/jclp.20373

Steiner, V., Pierce, L., Drahuschak, S., Nofziger, E., Buchman, D., & Szirony, T. (2008). Emotional support, physical help, and health of caregivers of stroke survivors. The Journal of neuroscience nursing: Journal of the American Association of Neuroscience Nurses, 40(1), 48. 10.1097/01376517-200802000-00008

Stroke Association. (2023). A carer’s guide to stroke. [Leaflet] https://stroke-org-prod.codeenigma.net/sites/default/files/2023-09/Stroke_-%20_A_Carers%20_Guide_Digital.pdf

Stroke Association (2019). Lived experience of stroke report. Chapter 3-Caring for a stroke survivor. https://www.stroke.org.uk/lived-experience-of-stroke-report/chapter-3-caring-for-a-stroke-survivor

Stroke Association. (2023). What we think about: The stroke workforce. Overworked and undervalued: building a stroke workforce for the future. [Leaflet]. https://www.stroke.org.uk/sites/default/files/new_pdfs_2019/our_policy_position/psp_stroke_workforce.pdf

Subgranon, R., & Lund, D. A. (2000). Maintaining caregiving at home: a culturally sensitive grounded theory of providing care in Thailand. Journal of Transcultural Nursing, 11(3), 166–173. 10.1177/104365960001100302

Taylor, G. H., Todman, J., & Broomfield, N. M. (2011). Post-stroke emotional adjustment: A modified social cognitive transition model. Neuropsychological Rehabilitation, 21(6), 808–824. 10.1080/09602011.2011.598403

Thomas, E. W. S., Dalton, J. E., Harden, M., Eastwood, A. J., & Parker, G. M. (2017). Updated meta-review of evidence on support for carers. Health Services and Delivery Research. 10.3310/hsdr05120

Thomas, J., & Harden, A. (2008). Methods for the thematic synthesis of qualitative research in systematic reviews. BMC Medical Research Methodology, 8(1), 1–10. 10.1186/1471-2288-8-45

Thompson, S. C. (1991). The search for meaning following a stroke. Basic and Applied Social Psychology, 12(1), 81–96. https://psycnet.apa.org/doi/10.1207/s15324834basp1201_6

Törnbom, K., Persson, H. C., Lundälv, J., & Sunnerhagen, K. S. (2017). Self-assessed physical, cognitive, and emotional impact of stroke at 1 month: the importance of stroke severity and participation. Journal of Stroke and Cerebrovascular Diseases, 26(1), 57–63. 10.1016/j.jstrokecerebrovasdis.2016.08.029

Tricco AC, Lillie E, Zarin W, O’Brien KK, Colquhoun H, Levac D, et al. PRISMA Extension for Scoping Reviews (PRISMAScR): Checklist and Explanation. Ann Intern Med. 2018;169:467–473. doi: 10.7326/M18-0850.

Tsiakiri, A., Vlotinou, P., Paschalidou, A., Konstantinidis, C., Christidi, F., Tsiptsios, D., … & Aggelousis, N. (2023). A Scoping Review on Coping Strategies and Quality of Life of Stroke Caregivers: Often Underestimated Variables in Stroke Recovery Process?. BioMed, 3(3), 349–368. 10.3390/biomed3030029

Van Puymbroeck, M., & Rittman, M. R. (2005). Quality-of-life predictors for caregivers at 1 and 6 months poststroke: Results of path analyses. Journal of Rehabilitation Research & Development, 42(6). 10.1682/jrrd.2005.01.0025

White, C. L., Lauzon, S., Yaffe, M. J., & Wood-Dauphinee, S. (2004). Toward a model of quality of life for family caregivers of stroke survivors. Quality of Life Research, 13, 625–638. 10.1023/b:qure.0000021312.37592.4f

Yeung, E. H., Szeto, A., Richardson, D., Lai, S. H., Lim, E., & Cameron, J. I. (2015). The experiences and needs of C hineseLJC anadian stroke survivors and family caregivers as they reLJintegrate into the community. Health & Social Care in the Community, 23(5), 523–531.

Zarzycki, M., Morrison, V., Bei, E., & Seddon, D. (2023). Cultural and societal motivations for being informal caregivers: a qualitative systematic review and meta-synthesis. Health Psychology Review, 17(2), 247–276. 10.1080/17437199.2022.2032259

Zarzycki, M., Seddon, D., Bei, E., Dekel, R., & Morrison, V. (2022). How culture shapes informal caregiver motivations: A meta-ethnographic review. Qualitative Health Research, 32(10), 1574–1589. 10.1177/10497323221110356

Zawawi, N. S. M., Aziz, N. A., Fisher, R., Ahmad, K., & Walker, M. F. (2020). The unmet needs of stroke survivors and stroke caregivers: a systematic narrative review. Journal of Stroke and Cerebrovascular Diseases, 29(8), 104875. 10.1016/j.jstrokecerebrovasdis.2020.104875

